# IgA1 hinge-region O-glycoform signatures associated with disease phase and kidney involvement in pediatric IgA vasculitis: a cross-sectional mass-spectrometry study

**DOI:** 10.64898/2026.08.21.26361008

**Authors:** Jerome Vialaret, Anne Filleron, Renaud Cezar, Manuela Pastore, Marc Fila, Christelle Reynes, Jana Kindermans, Adrien Schvartz, Thierry Chevallier, Pierre Corbeau, Christophe Hirtz, Tu-Anh Tran

**Author notes:** **Corresponding author:** Tu-Anh TRAN, MD, PhD Department of Pediatrics, Nîmes University Hospital, Centre Hospitalier Universitaire de Nîmes, Service de pédiatrie, Place du Pr R. Debré, 30029 Nîmes Cedex 9, France, Prof. Christophe Hirtz, LBPC–PPC, Université de Montpellier, IRMB, CHU de Montpellier, INM/INSERM, 80 avenue Augustin Fliche, 34295 Montpellier, France.

## Abstract

**Background:** IgA vasculitis (IgAV) is the most common systemic vasculitis in children, and its prognosis is largely determined by renal involvement (IgAV nephritis). No routine blood test identifies IgAV or stratifies the risk of nephritis, although aberrant O-glycosylation of the IgA1 hinge region is central to its pathogenesis. We developed a mass-spectrometry assay to profile IgA1 hinge O-glycoforms and define signatures of disease activity and renal involvement.

**Methods:** IgA was affinity-purified from 5 µL of plasma from 91 children (27 with acute IgAV, 26 in remission, and 38 age-matched healthy controls; 24 with and 29 without nephritis), trypsin-digested, and hinge-region O-glycopeptides were quantified by LC-MS. Sixty-nine glycoforms were normalized to a total-IgA1 tryptic peptide. Duplicate measurements showed good analytical repeatability, with a median coefficient of variation of 6%. Groups were compared using Mann–Whitney and Kruskal–Wallis tests (Benjamini– Hochberg FDR). Discrimination was assessed by ROC analysis and cross-validated logistic regression panels.

**Results:** Acute IgAV showed broad remodeling of the hinge glycoform profile (35 glycoforms differed with excellent discrimination (AUC 0.93–0.95) for the best ones), with an increase in low-sialylated, agalactosylated species and a decrease in complex sialylated species. The profile was normalized in remission (no glycoform differed from the controls). Two distinct renal patterns emerged: disease-associated glycoforms already altered without nephritis and renal-specific glycoforms altered only in nephritis (H2N2S1, H3N3S5, H3N4S4, and H4N4S3). A four-marker panel discriminated nephritis among IgAV children with a cross-validated AUC of 0.86 (IC 95 % 0.75–0.94).

**Conclusions:** A single mass-spectrometry assay, from a small blood volume, captures an IgAV-associated IgA1 hinge O-glycoform signature that normalizes in remission, together with a distinct renal involvement associated signature. These findings identify candidate IgA1 O-glycoform signatures associated with IgAV activity and documented renal involvement. Prospective longitudinal studies are required to determine whether the renal-associated panel can predict subsequent nephritis.

## BACKGROUND

IgA vasculitis (IgAV) (also known as Henoch–Schönlein purpura) is the most frequent systemic vasculitis in childhood, mainly between 4 and 8 years of age [1–3]. Boys are most often affected, with a sex ratio varying from 1.2 to 1.6 / 1. The annual incidence varies from country to country, from 13 to 20 per 100,000 children before the age of 17 [4–6]. It is characterized by a triad of purpura, arthritis or arthralgia, and abdominal pain. The diagnosis criteria were published in 2010 by the European League against Rheumatism (EULAR), Pediatric Rheumatology European Society (PRES), and Pediatric Rheumatology International Trials Organization (PRINTO) [7, 8]. The prognosis of IgAV essentially depends on the presence or absence of renal involvement or IgAV nephritis. This complication affects approximately 40% of all children [2, 9].

In 80% of cases, it causes micro-hematuria with mild nephrotic proteinuria; however, in the remaining 20%, it causes nephrotic proteinuria and/or acute renal failure. Nephritis leads to the risk of developing renal damage in 11 to 30% of cases [9–11]. Around 2-5% of children with IgAV nephritis progress to renal failure or end-stage renal disease.

Renal damage occurs between the third day of diagnosis and the first year of diagnosis. Thus, 91% of patients who develop renal damage do so within the first six weeks, 97% within the first six months, and 99% at one year [12]. 1-year surveillance is recommended. The initial symptomatology of nephritis is variable, and in most cases discreet (70-80% with isolated microhematuria +/- mild proteinuria) and generally predictive of renal prognosis. This long delay in monitoring and the absence of symptoms felt by the patient raise the problem of patients being lost to follow-up or with irregular follow-up, leading to sub-optimal management. Patients with moderate symptoms at presentation (microhematuria, mild proteinuria) show a poor prognosis in 15% of cases, compared with 41% for patients whose initial presentation is severe [13].

Despite well-established clinical patterns, diagnosis and prognostication remain challenging. Current best practice relies primarily on clinical classification criteria without a disease-defining blood test. The EULAR/PRINTO/PRES 2010 criteria—widely used in children—require purpura/petechiae (mandatory) plus at least one feature such as abdominal pain, arthritis/arthralgia, renal involvement, or biopsy evidence of IgA deposition. This disease is well known to pediatricians but less so to general practitioners, and mention of differential diagnoses is common. Currently, there are no specific biomarkers that have proven clinically useful for routine diagnosis, and incomplete/atypical presentations (for example, abdominal pain preceding purpura) can delay classification and broaden the differential diagnosis at first contact [8, 14]. These limitations directly impact clinical decision-making, including the need for intensified renal monitoring, early nephrology referral, and/or kidney biopsy. Furthermore, it is impossible to target children who are at risk of renal damage to optimize their monitoring.

IgA vasculitis is an IgA-dominant small-vessel disease. In IgAV nephritis, IgA-containing immune complexes accumulate in the glomerular mesangium and trigger inflammation. Pathogenic models overlap with IgA nephropathy and include increased production of galactose-deficient IgA1, anti-glycan autoantibodies, immune complex formation, and impaired clearance [10, 11, 15–17].

Human IgA1 carries clustered O-glycans in the hinge region. These glycans are initiated by N-acetylgalactosamine (GalNAc), extended by galactose, and capped by sialic acid. Reduced galactosylation can expose GalNAc-containing epitopes and promote the formation of nephritogenic immune complexes [11, 18–20].

At the molecular level, reduced galactosylation exposes GalNAc-containing epitopes in the IgA1 hinge region. These epitopes can be recognized by anti-glycan antibodies, promoting circulating immune-complex formation, mesangial deposition, and proinflammatory signaling [10, 19–21].

This glycosylation anomaly is found in variable proportions in various pathologies. In IgA nephritis (i.e., Berger’s disease), galactose-deficient serum IgA1 is associated with disease progression [22]. Thus, 77% of patients with IgA nephropathy have levels of poorly glycosylated IgA exceeding the mean observed in healthy individuals by more than two standard deviations [22]. In IgAV with renal involvement and without renal involvement, respectively 54% and 33% of these populations have elevated serum levels of poorly-glycosylated IgA; while only 6% of healthy subjects have elevated serum levels [23]. The detection of poorly glycosylated IgA in serum may therefore be associated with renal involvement, although it cannot be used to predict future nephritis [23].

Lectin-based studies have reported increased galactose-deficient IgA1 levels in IgA vasculitis nephritis, although the results vary across cohorts and assays. Elevated galactose-deficient IgA1 is not specific to IgAV nephritis and may also occur in IgAN or IgAV without nephritis. However, its association with renal severity and progression remains inconsistent [23–26]. Lectin-dependent immunoassays have been widely used to measure galactose-deficient IgA1; however, they have important limitations. The results depend on lectin specificity, neuraminidase treatment, standards, and assay calibration. Mass spectrometry provides direct compositional information and can resolve multiple hinge-region glycoforms in a single analysis [21, 23, 27].

Previous studies have established an association between abnormal IgA1 O-glycosylation and IgA vasculitis nephritis [15, 23, 24, 28–30]. Allen *et al*. [24] reported a lectin-defined abnormality restricted to patients with clinical nephritis. Nakazawa *et al.* [30] later used MALDI-TOF mass spectrometry in seven adults with end-stage IgA vasculitis nephritis and found reduced galactosylation after desialylation, with profiles similar to IgA nephropathy.

However, these studies did not provide a detailed, sialylation-preserving profile of a pediatric cohort. The adult MALDI-TOF study used desialylated glycopeptides and included transplant candidates with end-stage kidney disease. Therefore, it could not assess sialylation or early pediatric renal involvement.

Recent LC–MS studies have expanded IgA1 glycosylation analysis beyond the measurement of global galactose-deficient IgA1. Ohyama *et al.* [27] developed a workflow combining LC–HRMS, sequential deglycosylation, and EThcD fragmentation to quantify IgA1 hinge-region O-glycoforms and localize galactose-deficient sites. The study demonstrated extensive site-specific microheterogeneity, but was primarily methodological.

Clinical studies have subsequently identified disease-associated IgA1 glycosylation patterns in IgAN. Chen *et al.* [31] quantified 42 hinge-region O-glycopeptides from 8 µL of plasma and found reduced sialylation in IgA nephropathy, with an association between sialylation and eGFR. Zhang *et al*. [32] reported lower hinge-region GalNAc and galactose content in biopsy-proven IgA nephropathy than in other glomerular diseases or healthy controls. A panel combining these traits with circulating IgA levels achieved an AUC above 0.90.

In the present study, we therefore used LC–MS to characterize a broad panel of intact IgA1 hinge-region O-glycoform compositions in children with IgA vasculitis. We examined glycoform changes associated with acute disease, remission, and renal involvement. Our approach preserved the sialylation dimension and directly addressed whether renal involvement is associated with a distinct IgA1 glycoform signature.

## METHODS

### Study population

This study included three groups of subjects: A, patients in the acute phase of IgAV according to the EULAR/PRES/PRINTO 2010 classification (n=27); B, patients in remission of IgAV (n=26); and C, healthy controls (HC) (n=38) matched for age with patients. For IgA analysis of glycosylation, we included 27 patients in group A, including 13 with nephritis, and 26 patients in group B, including 11 with nephritis. Among the 11 nephritis cases, six were documented by renal biopsy. IgA Subjects were included between February 2015 and July 2017 in two French university hospitals. The subjects were between 3 and 18 years of age. The exclusion criteria were other autoimmune or inflammatory diseases and immunosuppressive treatment, biologics, or antibiotics during the previous 3 months.

The population and immunological characteristics have already been described [33]. For renal involvement, 45% of patients had proteinuria ≥ 30 mg/dl and/or hematuria ≥ 80 red cells/microliter) on dipstick. For the total population, IgAV patients were categorized as IgAV patients with biopsy-documented nephritis (IgAVNb) (n=7), nephritis with urine test diagnosis (IgAVNu) (n=17) (YES), or without nephritis (IgAVw) (n=29) (NO).

### Clinical samples

#### Sample collection and processing

The present study included 91 archived plasma samples collected within the prospective FOX-TREG study (ClinicalTrials.gov identifier: NCT02317133); a subset of these samples was also included in the ancillary FOXIGA-2020 study (ClinicalTrials.gov identifier: NCT04655378). Participants were recruited at Nîmes University Hospital and Montpellier University Hospital, France, between February 2015 and July 2017.

At inclusion, 10 mL of peripheral venous blood was collected into Vacutainer tubes containing sodium heparin as the anticoagulant. Blood samples were processed according to the routine pre-analytical procedures of the participating centers, generally within 2 h of venipuncture. Following plasma separation, samples were aliquoted and stored at −80 °C until release for the present study. None of the 91 plasma samples had undergone a freeze–thaw cycle before release. For the discovery analysis, sodium-heparin plasma samples from five patients with IgAV nephritis were analyzed on a timsTOF HT mass spectrometer.

#### Ethical approval

The FOX-TREG study and the subsequent use of the archived biological samples were approved by the CPP Sud Méditerranée III ethics committee (approval no. 2013.10.05) and by the relevant institutional review board under approval nos. 19.01.06 and 20.0061. The associated biological collection was declared under reference DC-2019-3463. Written informed consent was obtained from the participants’ parents or legal guardians, and age-appropriate information was provided to the children, whose assent was obtained whenever applicable.

#### Chemicals

Chemicals were purchased from ThermoFisher (Waltham, USA) (CaptureSelect, Pierce spin column), Sigma-Aldrich (Saint Louis, USA) DTT: DithioThreitol, IAA: iodoacetamide, Tris HCl, and guanidine hydrochloride, Biosolve (Dieuze, France) formic acid, acetonitrile, and water, Agilent (Santa Clara, USA) vials, Dutsher (Bernolsheim, France) PBS 10x, Merck (L’Isle-d’Abeau Chesnes, France) ammonium bicarbonate, VWR (Radnor, USA) NaCl, EDTA, and Promega (Promega, Madison, USA) trypsin.

#### Sample preparation for mass spectrometry analysis: Protein purification

The CaptureSelect IgA Affinity Matrix was used to purify human IgA. In a Pierce spin column with cap, 2 µL of CaptureSelect, 5 µL of plasma sample, 145 µL of PBS (1X), and 50 µL of incubation buffer (pH 7.4, 20 mM Tris HCl, 137 mM NaCl, 2 mM EDTA, 5 mM guanidine hydrochloride, 1% NP-40) were added and incubated at room temperature for 1 h at 750 rpm. The column was then washed with 400 µL of incubation buffer, 400 µL of PBS (1X), and 400 µL of H2O. Elution was performed with 100 µL of 100 mM Formic Acid, and samples were dried at 50°C.

After drying, the samples were resuspended in 30 µL of 50 mM ammonium bicarbonate (ABC) and 12.5% acetonitrile (ACN). Reduction was performed by adding 5 µL of 35 mM DTT and was incubated at 56°C for 30 min at 450 rpm. Samples were cooled down on ice and alkylation was performed by adding 5 µL of 125 mM IAA and incubating at 37°C during 30 minutes at 450 rpm. The IAA was neutralized with 5 µL of 35 mM DTT and digestion was performed by adding 1 µg of trypsin and incubating on 37°C overnight at 450 rpm. The digestion was stopped by adding 3 µL of formic acid.

The peptides were cleaned using C18 tips (Agilent Technologies, Santa Clara, USA). The C18 tips were primed with 70% ACN/0.1% FA, equilibrated with 0.1% FA, and the samples were loaded. The peptide clean-up included washes using 0.1% FA, followed by elution using 70% ACN/0.1% FA. The cleaned peptides were dried using a Speedvac (Labconco, Kansas, USA). Samples were resuspended in 20 µL of phase A (2% ACN / 0.1% FA / 97.9% water), mixed at 450 rpm during 10 minutes and stocked in appropriate vials.

#### Protein identification by LC-MS

Peptides were loaded onto Evotips Pure according to the manufacturer’s instructions. The peptide concentration was quantified using a NanodropOne Microvolume UV-Vis Spectrophotometer (reference ND-ONE-W, ThermoFisher Scientific) at 205 nm. A total of 400ng of peptides were desalted, and peptide separation was performed using an EvoSep One liquid chromatography system (EvoSep) [34] with a PepSep C18 column, 15 cm x 150µm, 1.5µm (Bruker Daltonics). Elution was carried out over 34-minutes gradient corresponding at 30 sample per day (SPD) using mobile phase A (0.1% FA in water; Biosolve) and mobile phase B (0.1% FA in acetonitrile; Biosolve). Peptides were analyzed using a trapped ion mobility spectrometry quadrupole time-of-flight mass spectrometer (timsTOF HT, Bruker Daltonics) equipped with a nanoelectrospray ion source (Captive spray, Bruker Daltonics) in positive-ion mode.

For glyco-PASEF method, the source parameters were as follows: capillary voltage, 1100 V; dry gas, 3.0 L/min; and dry temperature, 180 °C. MS1 and MS2 spectra were collected in the range of 100–3000 m/z with a mobility range of 0.75 to 1.40 1/K0. The accumulation and ramp times were 100 ms.

The collision energy was ramped by linear interpolation between 0.5 and 1.60 1/K0. The collision energy ranges was of 35.0–40.0 eV and 65.0–100.0 eV, respectively. A glycan-specific polygon was employed.

The upper boundary were 800.5m/z/1.05 1/K0; 1400.6m/z/1.40 1/K0; 1705.0m/z/1.40 1/K0; and for the lower boundary 802.6m/z/0.80 1/K0; 1702.8m/z/1.10 1/K0. Ten PASEF ramps were used with a range of charge state selection (2-6) and an intensity threshold of 2500. Three ions of the Agilent ESI-Low Tuning Mix were used to calibrate the ion mobility dimension (m/z [Th], 1/K0 [Th]: 622.0289, 0.9848; 922.0097, 1.1895; 1221,9006, 1.3934).

Raw LC–MS/MS data files (Bruker. d format (DDA acquisition) were processed in FragPipe v22.0 using MSFragger v4.1 and Philosopher v5.1.1.

MSFragger search was performed in labile mode for O-glyco search with 20ppm mass tolerance. The enzymatic specificity was semi-enzymatic specificity for trypsin with a maximum of two missed cleavages. The static cysteine alkylation was set to +57.02146 Da. Variable modifications included oxidation of Met (+15.9949 Da) and N-terminal acetylation (+42.0106 Da). The identification results were filtered sequentially with a 1% protein-level FDR. PTM-Shepherd v3.0.0 was used for glycan assignment. For spectral-library creation, the five individual MSFragger pepXML files were manually imported into Skyline/BiblioSpec using an expectation-type PSM score threshold of 0.01.

#### Targeted profiling of IgA1 hinge-region O-glycopeptides

One microliter of each sample was injected into nanoElute (Bruker Daltonics, Massachusetts, USA). NanoFlow LC was coupled to Q-TOF MS instrument (Impact II, Bruker Daltonics, Massachusetts, USA) through captive spray ion source (1200V, dry gas: 3l/min at 150°C) operating with nanobooster (0.2 Bar of Nitrogen boiling in acetonitrile). Using the LC, samples were desalted and pre-concentrated on-line on a PepMap u-precolumn (300 µm x 5 mm, C18 PepMap 100, 5 µm, 100 Angström, ThermoFisher, Massachusetts, USA). To perform separation, peptides were transferred to an analytical column (75 µm × 500 mm; Acclaim Pepmap RSLC, C18, 2µm, 100 Angström, ThermoFisher, Massachusetts, USA). A gradient consisting of 5-26% B for 192 min and 90% B for 10 min (A = 0.1% formic acid, 2% acetonitrile in water; B = 0.1% formic acid in acetonitrile) at 400 nL/min, 50°C was used to elute peptides from the reverse-phase column.

Glycopeptides were monitored using LC-MS profiling. A mass range of 490–2500 m/z was scanned at 1.0 Hz with a lock-mass as internal calibrator (m/z 1222, Hexakis “1H, 1H, 4H-hexafluorobutyloxy’’ phosphazine, Agilent Technologies, Santa Clara, USA).

#### Cohort quantification

Skyline 21.1 was used to process the raw data. Based on the identified peptide list and retention time determined by the identification step, the masses were extracted from the raw data from the profile analysis. Transition Settings were adjusted to extract Precursor Ion with a positive charge of 4 and 5 in the mass range of 500-3000 m/z. The m/z mass tolerance was 0.05, and the retention time tolerance was 2 min. All peaks for precursor charges 4+ and 5+ were checked, integrated, and exported as an Excel file for supplementary analysis. A feature was retained when its isotope envelope was consistent with the expected precursor charge and monoisotopic assignment, its measured mass was within the predefined extraction tolerance, no relevant coeluting interference was observed, and the chromatographic signal was clearly distinguishable from local noise and reproducibly integrated across technical duplicates.

#### Compounds monitored

DASGVTFTWTPSSGK, H0N1S0, H1N1S0, H1N1S1, H1N1S2, H1N2S0, H1N2S1, H1N3S0, H1N3S1, H1N4S1, H2N2S0, H2N2S1, H2N2S2, H2N3S0, H2N3S1, H2N3S2, H2N4S0, H2N4S1, H2N4S2, H2N5S0, H2N5S1, H2N5S2, H3N3S0, H3N3S1, H3N3S2, H3N3S3, H3N3S4, H3N3S5, H3N4S0, H3N4S1, H3N4S2, H3N4S3, H3N4S4, H3N5S0, H3N5S1, H3N5S2, H3N5S3, H3N5S4, H3N6S0, H3N6S1, H3N6S2, H3N6S3, H4N4S0, H4N4S1, H4N4S2, H4N4S3, H4N4S4, H4N4S5, H4N5S0, H4N5S1, H4N5S2, H4N5S3, H4N5S4, H4N5S5, H4N6S0, H4N6S1, H4N6S2, H4N6S3, H4N6S4, H5N5S0, H5N5S1, H5N5S2, H5N5S3, H5N5S4, H5N5S5, H5N6S2, H5N6S3, H5N6S4, H6N6S4, DASGVTFTWTPSSGK correspond to total Iga, HYTNP..HPR is the unglycosylated part of the hinge region.

Derived traits summarized the weighted hexose, HexNAc, and sialic-acid content; the abundance of low-sialylated and highly sialylated compositions; and the balance between hexose and HexNAc content.

HYT_H_N, HYT_LowSsum, HYT_NbH: Intensity multiply by number of galactoses (H), HYT_NbN: Intensity multiply by number of N-Acetyl-galactosamines (N), HYT_NbS: Intensity multiply by number of sialic acid (S), HYT_NsupH, HYT_S_H, HYT_SinfH, HYT_SsupH, HYT_highSsum.

HYTNP..HPR, HYT_H_N: sum of intensities multiplied by galactoses per N-acetyl-galactosamine fraction, HYT_LowSsum: Relative intensity sum of structures with 0, 1, and 2 S, HYT_NbH, HYT_NbN, HYT_NbS, HYT_NsupH: Relative intensity sum of structures in which the number of N-acetyl-galactosamines exceeds the number of galactoses, HYT_S_H: sum of intensities multiplied by sialic acid per galactoses fraction, HYT_SinfH: Relative intensity sum of structures in which the number of sialic acids is lower than the number of galactoses, HYT_SsupH: Relative intensity sum of structures in which the number of sialic acids exceeds the number of galactoses, HYT_highSsum: Relative intensity sum of structures with 4, 5, and 6 S.

Structure–abundance relationship. To test whether the direction and magnitude of the acute-phase differential abundance of IgA1 hinge-region O-glycopeptides depended on glycan composition, each glycan was decomposed from its HxNySz identifier into the number of hexose (H), N-acetylhexosamine (N), and sialic acid (S) residues. For each glycan, enrichment in the acute phase was defined as the difference in mean log2 abundance between acute IgAV and healthy controls (mean log2[acute] − mean log2[control]; positive values indicated higher abundance in acute IgAV). Across the 69 glycans, this enrichment value was correlated with the number of sialic acid and HexNAc residues using Spearman’s rank correlation coefficient (ρ). This comparison was performed for all combinations.

#### Statistical analysis

Statistical analyses were performed using RStudio (version 2025.09.01). Each participant contributed one biological sample and was assigned to a clinical group. Technical LC–MS duplicates were averaged before inferential statistical analysis and were not treated as independent observations. Glycopeptide abundance was normalized to the IgA1 proteotypic peptide DASGVTFTWTPSSGK and log2-transformed.

The single missing value (glycan H2N4S2 in sample C01P026) was imputed with the global median of that glycan across all samples (5873.4, raw normalized-area scale). Overall differences among acute IgAV, IgAV in remission, and healthy controls were assessed using the Kruskal–Wallis test. Pairwise comparisons were performed using the Mann–Whitney U test for acute IgAV versus remission, acute IgAV versus healthy controls, and remission versus healthy control groups. Renal status comparisons included healthy controls versus IgAV without nephritis, healthy controls versus IgAV with nephritis, and IgAV without versus with nephritis. P values were adjusted for multiple testing across the 69 quantified glycoforms within each comparison using the Benjamini–Hochberg procedure. An FDR-adjusted P-value < 0.05 was considered statistically significant. The discriminative performance was assessed using receiver operating characteristic curves and summarized by the area under the curve. Internal validation and feature-selection stability in SupData 1.

To test whether disease activity confounded the renal-associated changes, patients were cross-classified into four subgroups (acute/remission x nephritis/no-nephritis) and the four candidate glycoforms were compared with two-sided Mann-Whitney tests stratified by activity (nephritis vs no-nephritis within acute, and within remission) and stratified by renal status (acute vs remission within nephritis, and within no-nephritis); p-values were adjusted by the Benjamini-Hochberg procedure across the four markers within each comparison. In addition, a two-way linear model of log2 abundance including activity, renal status and their interaction was fitted for each marker, and the interaction term was used to test whether the magnitude of the renal effect differed between acute and remission.

## RESULTS

### Context

Plasma IgA was purified from 5 µL of sample, digested with trypsin, and analyzed using liquid chromatography–mass spectrometry (Figure 1). The normalized areas of glycopeptides were compared between the groups. The population has already been described by Filleron et al. 2024 [33] (Figure 2C). The population of patients with the same disease could be separated. The cohort comprised 27 acute IgA vasculitis samples (A), 26 remission samples (B), and 38 healthy control samples (C). The groups were comparable in terms of age and sex (Table 1). IgA vasculitis samples were also classified according to renal involvement: IgAV with nephritis (n=24) (YES) or without nephritis (n=29) (NO).

**Figure 1.**
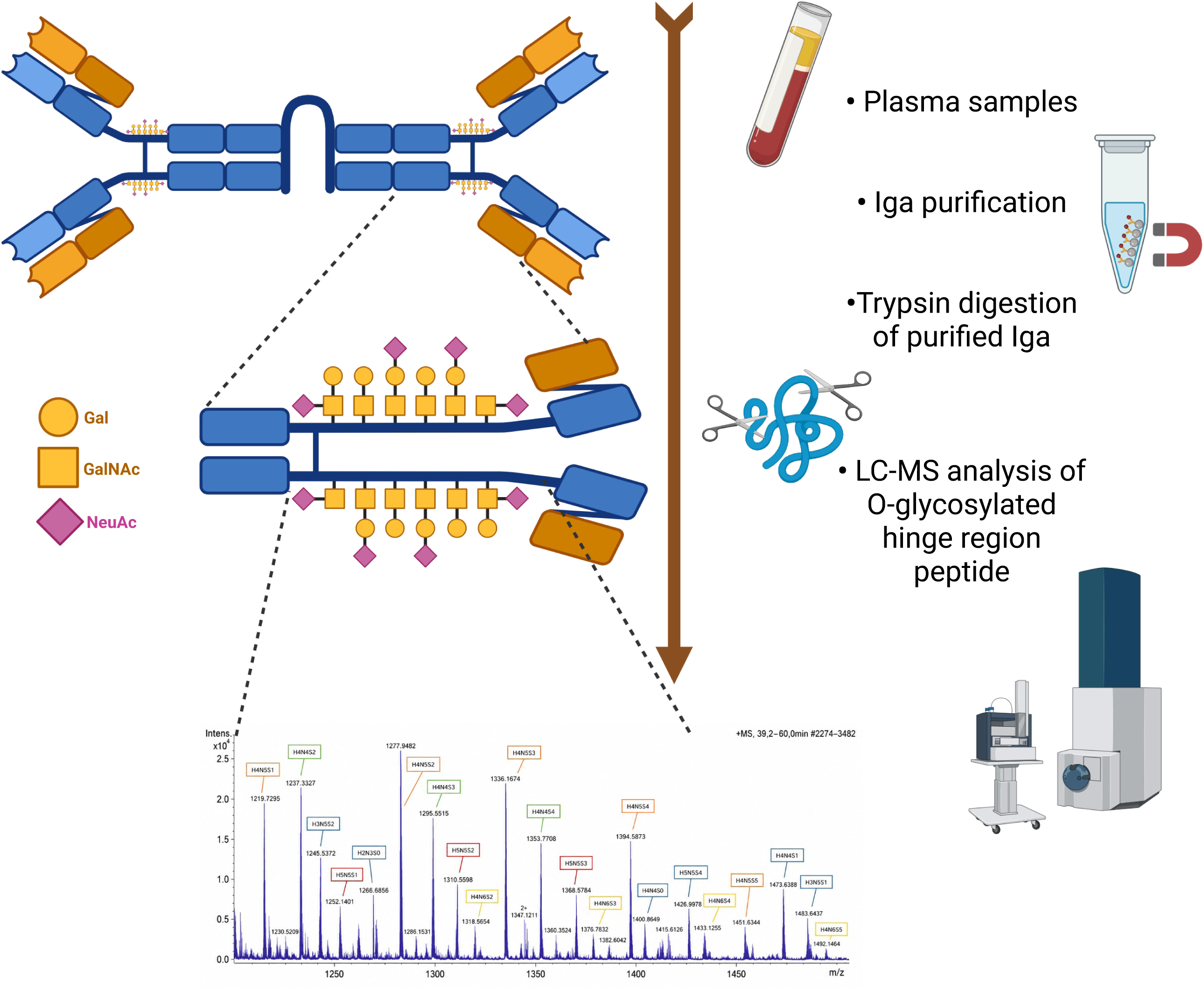
Workflow for IgA1 glycoform preparation and mass spectrometric analysis. IgA1 was isolated from plasma samples using affinity-based enrichment and subsequently subjected to enzymatic digestion. The resulting hinge-region glycopeptides, carrying variable combinations of galactose (Gal), N-acetylgalactosamine (GalNAc), and N-acetylneuraminic acid (Neu5Ac), were analyzed by mass spectrometry to characterize and quantify IgA1 O-glycoforms.

**Figure 2.**
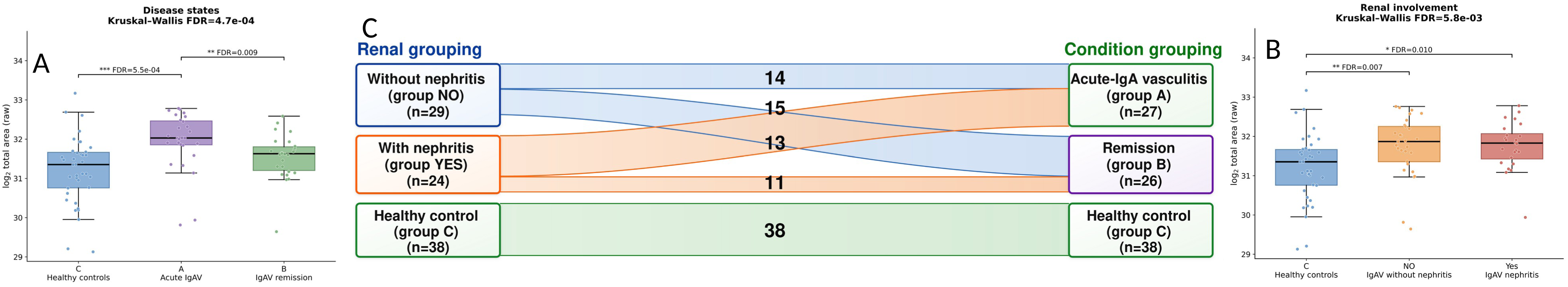
Distribution of study participants and total IgA1 according to renal involvement and clinical status. C: The alluvial diagram shows the distribution of pediatric IgA vasculitis patients with (YES, n = 24) or without nephritis (NO, n = 29) between the acute phase (A, n = 27) and remission (group B, n = 26). Healthy controls constituted group C (n = 38). Numbers within the connecting bands indicate the number of participants in each subgroup. A, B: Total IgA1 marker peptide DASGVTFTWTPSSGK across groups. Abundance of the non-glycosylated IgA1 tryptic peptide DASGVTFTWTPSSGK, used as a proxy for total IgA1, shown as log2 total (raw) peak area per sample. A: disease states - healthy controls (C, n = 38), acute-phase IgA vasculitis (A, n = 27) and IgAV in remission (B, n = 26). B: renal involvement - healthy controls (C, n = 38), IgAV without nephritis (NO, n = 29) and IgAV with nephritis (YES, n = 24).

**Table 1:** Demographic and clinical characteristics of the populations under study.

| Group | A | B | C |
| --- | --- | --- | --- |
| Number of subjects | 27 | 26 | 38 |
| Sex (M/F) | 12/15 | 15/11 | 22/16 |
| Age (years) | 6.1 (4.4; 7.2) | 6.6 (5.4; 8.0) | 6.5 (4.8; 9.5) |
| Weight (kg) | 18 (16.5; 25.5) | 22 (18.3; 31) | 21.5 (18; 34) |
| Height (cm) | 110 (102; 119.5) | 119 (113.3; 130.3) | 120 (108.3; 138.8) |
| Body mass index (kg/m<sup>2</sup>) | 16.02 (14.09; 17.61) | 15.67 (14.65; 16.60) | 15.75 (15.03; 17.35) |
| Systolic blood pressure (mmHg) | 101 (96.5; 112) | 96 (92; 106) | 100 (93; 110.3) |
| Cutaneous involvement | 27 | 26 | - |
| Abdominal involvement | 17 | 18 | - |
| Articular involvement | 25 | 26 | - |
| Renal involvement |  |  | - |
| Proteinuria ≥ 1+ and/or Hematuria≥2+ | 13 | 11 |  |
| Biopsy documented nephritis | 1 | 6 |  |
| Genital involvement (orchitis) | 3 | 3 | - |

### IgA1 identification and normalization

IgA1 is the protein of interest for studying O-glycosylation regulation in the hinge region. IgA1 was identified with six peptides corresponding to 8-51; 52-81; 154-168; 201-212; 283-299; 307-398. The correlation between peptides were between 0.7816 [154–168] and 0.6749 [307–398] indicating a good correlation between the peptides (only exception was the long N termini peptide [8–51] with 0.537). Peptide 154-168 (DASGVTFTWTPSSGK) was used to quantify IgA1.

Relative circulating IgA1 abundance differed between healthy controls, acute IgAV, and IgAV remission (Kruskal–Wallis, BH-adjusted q = 4.7 × 10⁻⁴). Abundance was higher in acute IgAV than in healthy controls (q = 5.5 × 10⁻⁴) and remission (q = 0.009), whereas remission and healthy controls did not differ significantly (Figure 2A). IgA1 abundance also differed between healthy controls and IgAV subgroups defined by renal status (q = 5.8 × 10⁻s). Both IgAV patients without nephritis (q = 0.007) and those with nephritis (q = 0.010) had higher IgA1 abundance than controls, but no difference was observed between the two IgAV subgroups (Figure 2B).

### IgA1 hinge-region O-glycoform coverage

A total of 85 glycosylation masses were identified in the hinge region peptide. Supplementary Table 1 presents the different identified sites. Redundancy was removed by retaining the better Hyperscore. The most intense glycopeptides presented more than 200 PSMs, such as H4N4S2, H4N5S4, H4N5S2, H4N4S4, H4N5S1, and H4N5S3 (Sup Figure 1A). CID fragmentation was not optimal for confident site localization, especially on this long and Ser/Thr rich peptide but Fragpipe provided probabilistic localization scores. Positions S17/T26/T18/T29/T21/S23 are the sites, in decreasing order, with the highest number of assigned glycosites (Sup Figure 1E).

In a sample cohort of 91 samples, careful inspection was performed on the data, and consequently, 69 glycosylation were used to perform a quantitative comparison. Removed glycopeptides presented noise, were too broad, interfered, or had low precision peaks.

To explore the glycan diversity inside the cohort, glycan abundances were expressed as percentage of the total HNS signal, and cohort summaries were reported as the median across patients. The profile was dominated by a limited set of highly sialylated species, with H4N5S2, H4N5S3, H4N4S2, and H4N4S3 each contributing ∼6–7% (median) of the total HNS pool (Sup Figure 1A). The dataset displayed a wide dynamic range, with median glycan abundances spanning ∼0.03% to ∼7.41% (∼247×), and individual measurements spanning ∼0.003% to ∼13.94% (∼4,383×), with the largest relative variability observed among low-abundance glycans.

Overall, sialylated glycans represented a median of ∼91.6% of the HNS signal (S1–S2: 49.3%; S3+: 42.2%) (Sup Figure 1C). The compositional imbalance between N and H is a descriptor of the galactosylation-deficient index (GDI) when the GDI is below 1. The signal was primarily distributed across N−H = 0–1 (∼83.9%), with a smaller contribution from N−H ≥2 (Sup Figure 1B). The complexity of the sugar, based on HexNAc composition, is overwhelmingly carried by N4–N6 species (Sup Figure 1D)

A total of 91 samples were analyzed in duplicate, and the median coefficient of variation was 6.0% (4.0-10.6%) (Sup Table 2).

### Disease-phase signature

IgA1 hinge-region O-glycoforms were quantified in plasma, and three clinical groups were compared: A (patients in acute phase of IgA vasculitis (IgAV) (n = 27); B (patients in remission of IgAV (n = 26); C (healthy controls (n = 38). A three-group non-parametric comparison (Kruskal–Wallis test) revealed widespread differences across disease stages, with 44 glycoforms remaining significant after FDR correction (FDR ≤ 0.0303). The strongest overall group effects were observed for H3N3S0 (H = 42.296; η² = 0.458), H5N5S0 (H = 42.222; η² = 0.457), and H4N4S0 (H = 40.881; η² = 0.442).

Thirty glycoforms differed between acute IgA vasculitis and remission (Figure 3). Low-sialylated glycoforms were highly enriched in acute IgA vasculitis. H3N3S0 (log2 fold change, 1.93; FDR, 1.0 × 10⁻⁶; AUC, 0.95), H5N5S0 (log2 fold change, 2.11; FDR, 1.0 × 10⁻⁶; AUC, 0.95), H4N4S0 (log2 fold change, 1.57; FDR, 1.0 × 10⁻⁶; AUC, 0.94), and H3N3S1 (log2 fold change, 1.29; FDR, 1.0 × 10⁻⁶; AUC, 0.93) showed the strongest separation. Collectively, acute-phase signatures were characterized by compositions with approximately balanced Hex/HexNAc counts (H≈N) and low sialylation (S0–S1), suggesting a shift toward less sialylated, compositionally “balanced” hinge O-glycoforms during acute inflammation. Conversely, remission showed higher levels of sialylated HexNAc-rich compositions. These included H3N5S3 (log2FC −1.10, FDR 2.55×10⁻^4^, AUC 0.83), H3N5S2 (log2FC −0.80, FDR 5.43×10⁻⁴, AUC 0.81), H3N6S3 (log2FC −1.09, FDR 9.04×10⁻⁴, AUC 0.80), and H3N5S4 (log2FC −1.16, FDR 1.09×10⁻⁴, AUC 0.79). These glycoforms typically show lower H/N ratios and higher sialylation, consistent with a compositional shift toward more sialylated and HexNAc-rich hinge O-glycoform patterns in remission. Glycoforms decreased in acute IgAV were enriched for higher sialylation and HexNAc-rich compositions.

**Figure 3.**
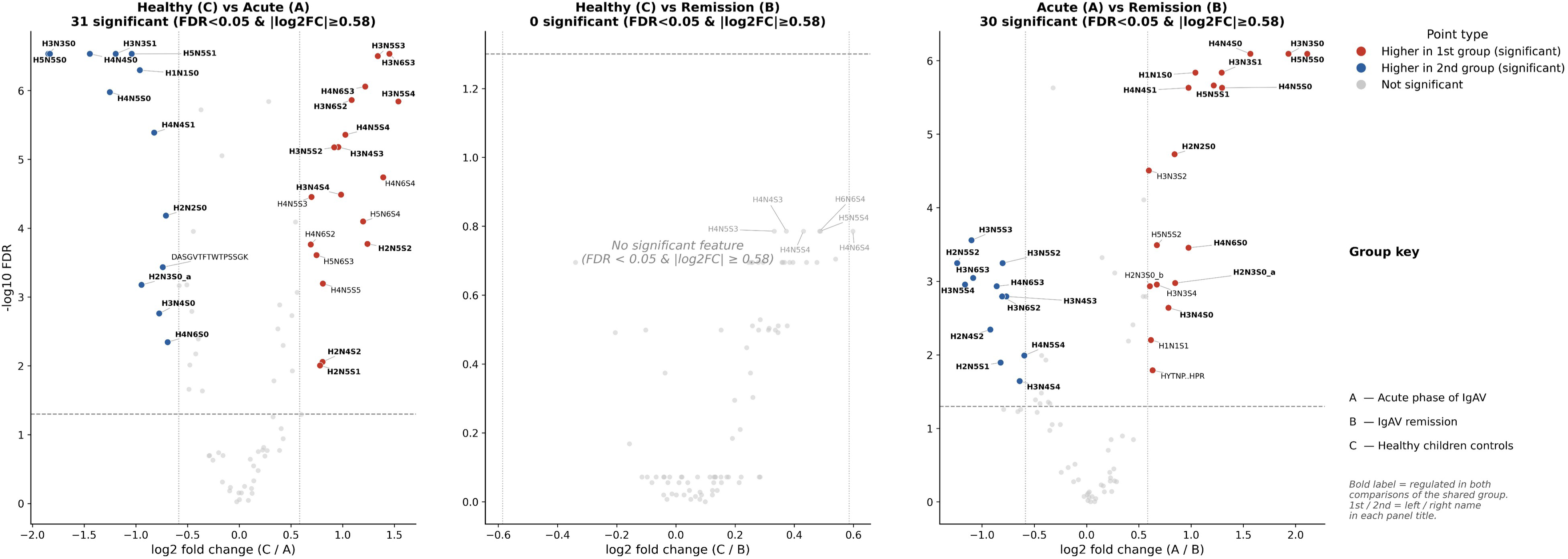
Volcano plots - IgAV disease states (C / A / B). Healthy controls (C), acute-phase IgAV (A) and IgAV in remission (B). Panels: C vs A (31 regulated), C vs B (0 regulated), A vs B (30 regulated). Acute IgAV (A) carries a strong, symmetric signature versus both healthy controls and remission, whereas remission (B) is statistically indistinguishable from healthy controls. For reference, the six most extreme (but non-significant) features of the C-vs-B panel are labeled in pale grey: H4N6S4 (log2FC 0.598, P = 0.0040, AUC 0.714), H4N5S3 (0.332, 0.0089, 0.694), H4N5S4 (0.431, 0.0096, 0.692), H5N5S4 (0.488, 0.0117, 0.687), H4N4S3 (0.373, 0.0121, 0.686) and H6N6S4 (0.485, 0.0121, 0.686); P is the raw Mann-Whitney p-value (all share FDR = 0.16, i.e. not significant). Bold labels (n = 24) are the features regulated in both C-vs-A and A-vs-B - the robust acute-phase signature; all are glycans, dominated by an increase in simple asialo/agalacto species (e.g. H3N3S0, H5N5S0, H4N4S0) and a decrease in complex sialylated glycans (e.g. H3N6S3, H3N5S3, H4N6S3). The number of regulated compounds was determined using FDR < 0.05 and |log2FC| ≥ 0.585.

Thirty-one glycoforms differed between acute IgA vasculitis and healthy controls (FDR <0.05, |log2FC| ≥0,58). The acute-versus-control profile strongly overlapped with the acute-versus-remission profile. This concordance supports a robust acute IgA vasculitis signature rather than a contrast-specific result.

No individual glycoform differed between remission and healthy controls after FDR correction (Figure 3). Six highly sialylated glycoforms showed nominal differences between controls (C) and remission (B) groups: H4N6S4 (log2FC, 0.598; P = 0.0040; AUC, 0.714), H4N5S3 (log2FC, 0.332; P = 0.0089; AUC, 0.694), H4N5S4 (log2FC, 0.431; P = 0.0096; AUC, 0.692), H5N5S4 (log2FC, 0.488; P = 0.0117; AUC, 0.687), H4N4S3 (log2FC, 0.373; P = 0.0121; AUC, 0.686), and H6N6S4 (log2FC, 0.485; P = 0.0121; AUC, 0.686). All were more abundant in controls than in remission, but none remained significant after FDR correction. These glycoforms were highly sialylated (S3–S4), and several were relatively HexNAc-rich.

### Disease-phase-associated IgA1 O-glycoform remodeling

Acute IgAV was associated with broad and coordinated remodeling of the IgA1 hinge region O-glycoform profile. Thirty-five glycoforms were differentially abundant in the same direction when comparing acute IgAV with either remission or healthy controls (Figure 3). Glycoforms enriched in acute disease were predominantly non-sialylated or monosialylated, including H3N3S0, H5N5S0, H4N4S0, and H3N3S1. In contrast, glycoforms depleted in acute IgAV were more sialylated and generally contained more HexNAc residues. Across the 69 quantified glycoforms, the acute-versus-control log2 fold change, defined as the mean log2 abundance in acute IgAV minus that in healthy controls, was strongly inversely associated with sialic acid count (Spearman ρ = −0.69, nominal p = 4 × 10⁻¹¹) and more moderately with HexNAc count (ρ = −0.46, nominal p = 7 × 10⁻⁵), but not with hexose count (ρ = −0.12, p = 0.33) (Figure 6). Acute-enriched glycoforms contained a mean of 0.25 sialic-acid residues, and 75% were non-sialylated, whereas control-enriched glycoforms contained a mean of 2.9 sialic-acid residues and none were non-sialylated.

The derived glycosylation traits reproduced the same low-sialylation axis but did not improve discrimination beyond individual glycoforms.

In the remission-versus-control comparison, the relationship between the fold change and sialic acid count remained directionally similar (ρ = −0.66, nominal p = 5 × 10⁻¹⁰), but the dispersion of fold changes was approximately fourfold lower than that in acute disease. Five highly sialylated glycoforms showed nominal differences between remission and control groups, with AUCs of 0.686–0.694; however, none remained significant after FDR correction. Therefore, the remission group showed a profile similar to that of the healthy controls. Because the acute and remission groups comprised different children, this cross-sectional pattern is consistent with near-normalization at the group level but does not demonstrate within-patient normalization.

### Renal-involvement signature

Global three-group (Controls [C, n=38], IgAV without renal involvement [NO, n=29], and IgAV with renal involvement [YES, n=24]) comparisons identified a subset of biomarkers whose distributions differed significantly according to disease status after correction for multiple testing. A three-group non-parametric comparison (Kruskal–Wallis test) showed widespread differences across disease stages with 32 glycoforms.

Compared with healthy controls, IgA vasculitis with renal involvement showed 32 significantly different glycoforms after FDR correction. Twenty-six were less abundant and six were more abundant in the renal-involvement group. H3N5S4 (log2 fold change, −1.472; FDR, 2.4 × 10⁻⁵; AUC, 0.89) and H3N4S4 (log2 fold change, −1.162; FDR, 2.4 × 10⁻⁵; AUC, 0.88) provided the strongest separation from healthy controls.

Five glycoforms differed between the controls and patients with IgA vasculitis without renal involvement (Figure 4). The best single-marker AUCs were 0.72–0.74. Several low-sialylated glycoforms were also increased in IgAV without renal involvement, with AUCs of 0.684–0.739.

**Figure 4.**
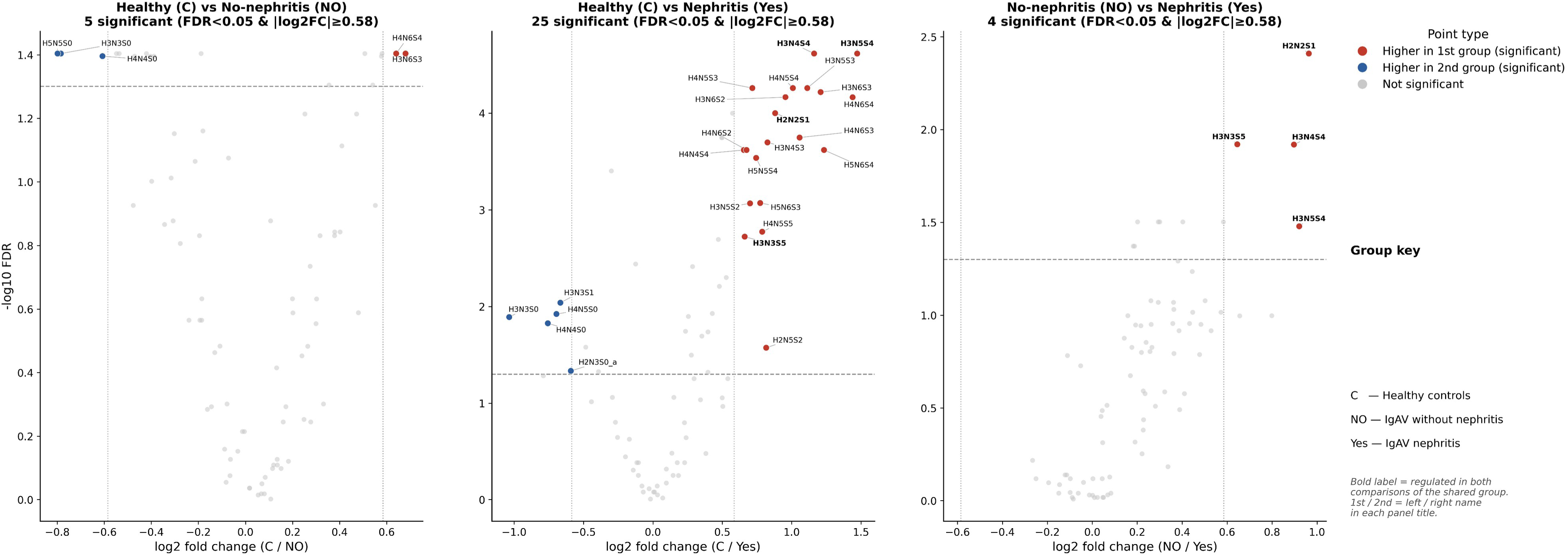
Volcano plots - renal involvement in IgAV (C / NO / YES). Healthy controls (C), IgAV without nephritis (NO) and IgAV with nephritis (YES). Panels: C vs NO (5 regulated), C vs YES (25 regulated), NO vs YES (4 regulated). The nephritis group (YES) shows the strongest shift; NO differs only modestly from controls. Bold labels are the features regulated in both C-vs-YES and NO-vs-YES - the nephritis-consistent signature, which resolves to exactly four O-glycoforms: H2N2S1, H3N3S5, H3N4S4 and H4N4S3. The number of regulated compounds was determined using FDR < 0.05 and |log2FC| ≥ 0.585.

**Figure 5.**
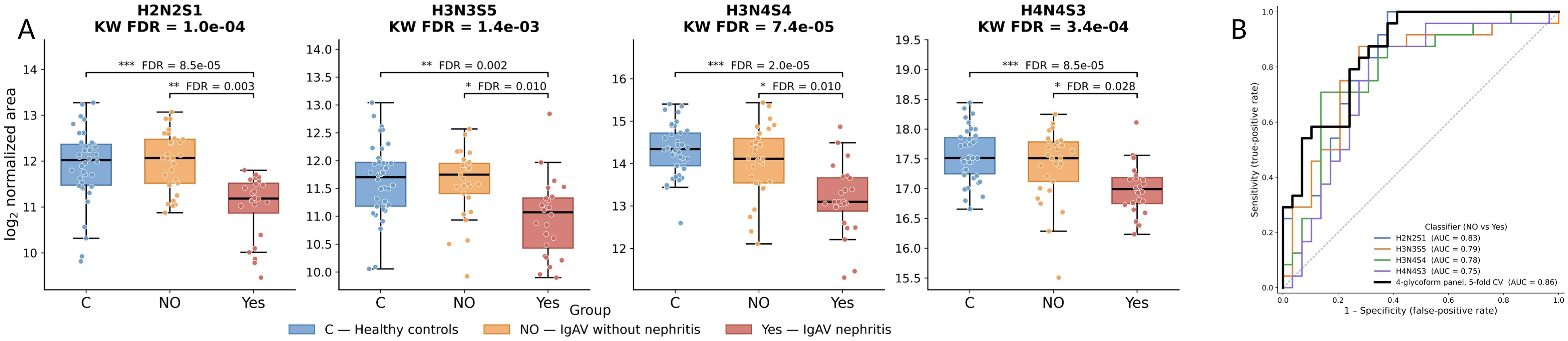
Renal-involvement-associated IgA1 O-glycoform signature. A: Renal-involvement-associated IgA1 O-glycoform signature: abundance across renal involvement. Distribution of four IgA1 O-glycoforms (H2N2S1, H3N3S5, H3N4S4, H4N4S3; log2 normalized area) in healthy controls (C, n = 38), IgA vasculitis without nephritis (NO, n = 29) and IgA vasculitis with nephritis (YES, n = 24). B: Renal-involvement-associated IgA1 O-glycoform signature: discrimination of IgAV nephritis. Receiver-operating-characteristic (ROC) curves for classifying IgA vasculitis patients with versus without nephritis (NO vs YES; positive class = nephritis, n = 24 vs n = 29). Coloured curves show each single O-glycoform used as a univariate classifier (area under the curve, AUC, in the legend). The black curve is a multivariable panel combining all four glycoforms in a logistic-regression model, evaluated by stratified 5-fold cross-validation (CV AUC). The dashed diagonal denotes chance performance.

### Renal-involvement IgA1 O-glycoform remodeling

Renal status analyses distinguished a broad IgAV-associated structural axis from a more restricted renal involvement-associated signature. Compared with healthy controls, 25 glycoforms differed in IgAV with nephritis, and 5 differed in IgAV without nephritis.

In both IgAV groups, glycoform-level log2 fold changes relative to controls were strongly inversely associated with sialic acid count: ρ = −0.73 for IgAV with nephritis versus controls (nominal p = 8 × 10⁻¹s) and ρ = −0.76 for IgAV without nephritis versus controls (nominal p = 4 × 10⁻¹⁴) (Figure 6A,B). This similar relationship in both groups indicates that reduced sialylation is a general IgAV-associated compositional axis rather than a renal-specific feature of IgAV. Nevertheless, the same relationship remained detectable in the direct comparison between IgAV with and without nephritis (ρ = −0.58, nominal p = 2 × 10⁻⁷), indicating an additional relative depletion of more highly sialylated glycoforms in renal involvement cases.

**Figure 6.**
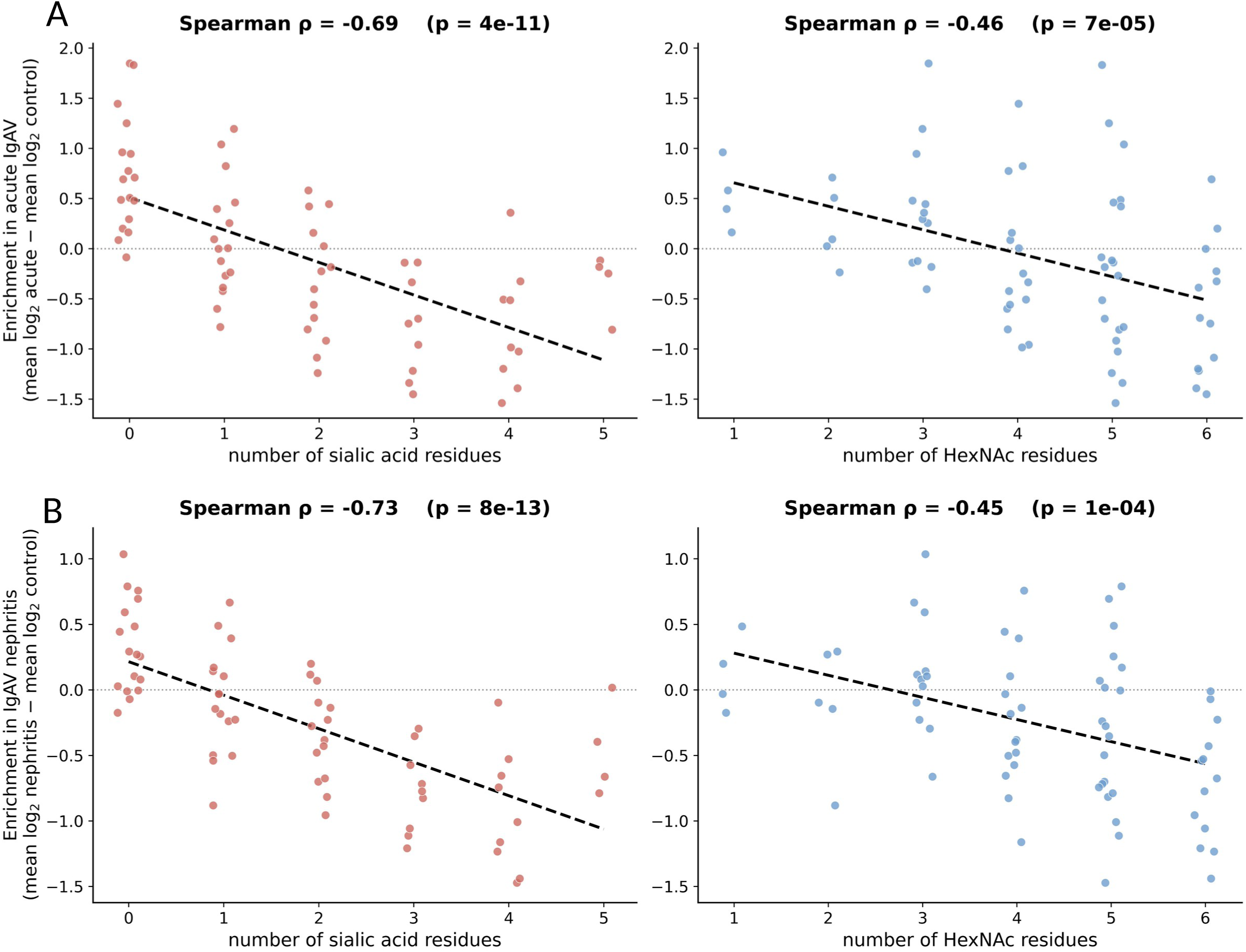
Relationship between IgA1 hinge-region O-glycopeptide composition and disease-associated abundance changes. A: Association between glycan composition and acute-phase differential abundance of IgA1 hinge-region O-glycopeptides. For each of the 69 glycans, enrichment in acute IgAV was computed as the difference in mean log2 normalized area between acute IgAV patients (A) and healthy controls (mean log2 acute - mean log2 control). Enrichment is plotted against the number of sialic-acid residues (left) and the number of HexNAc residues (right) in the glycan composition (HxNxSx). Each point is one glycan (x jittered for visibility); the dashed line is an ordinary-least-squares trend and the dotted line marks zero enrichment (no change vs control). Association was quantified by Spearman rank correlation (rho, p above each panel). Enrichment decreases with increasing sialylation (rho = −0.69, p = 4e-11) and, more weakly, with increasing HexNAc content (rho = −0.46, p = 7e-05): the more sialic acids or HexNAc a glycan carries, the more it is reduced in acute IgAV. B: Association between glycan composition and nephritis-associated differential abundance of IgA1 hinge region O-glycopeptides. As in Panel A, but enrichment is computed between IgAV patients with nephritis (YES) and healthy controls (mean log2 nephritis - mean log2 control), plotted against the number of sialic-acid (left) and HexNAc (right) residues. The negative composition dependence is even stronger for sialylation than in the acute-phase contrast (rho = −0.73, p = 8e-13) and comparable for HexNAc content (rho = −0.45, p = 1e-04), indicating that highly sialylated / more branched glycans are preferentially depleted in IgAV nephritis.

The clinically relevant comparison was IgA vasculitis with versus without renal involvement. Four glycoforms were significantly less abundant in the renal-involvement group: H2N2S1 (log2 fold change, −0.964; FDR, 0.00331; AUC, 0.83), H3N3S5 (log2 fold change, −0.645; FDR, 0.0102; AUC, 0.79), H3N4S4 (log2 fold change, −0.897; FDR, 0.0103; AUC, 0.78), and H4N4S3 (log2 fold change, −0.402; FDR, 0.0278; AUC, 0.75). A combined z-score reached an AUC of 0.86 (IC 95 % 0.75–0.94), exceeding the performance of each individual glycoform.

Because the primary nephritis comparison pooled patients sampled during the acute phase and remission, we performed a post hoc sensitivity analysis to determine whether disease phase influenced the renal-associated signature. Within the acute-phase group, all four glycoforms were less abundant in patients with nephritis after Benjamini–Hochberg correction across the four markers examined (FDR range, 0.024–0.039). The same direction of association was observed in remission (FDR range, 0.0018– 0.029). Conversely, after stratification by renal status, none of the four markers showed a detectable difference between the acute and remission groups, either among patients with nephritis or among those without nephritis. In two-factor models, no phase-by-renal-status interaction was detected for any marker (interaction FDR range, 0.36–0.76), and the renal-associated effect estimates were directionally consistent and of broadly comparable magnitude in both disease phases (Sup Figure 2). These findings confirm that disease phase does not account for the overall renal-associated signature, although the subgroup analyses had limited statistical power.

## DISCUSSION

This study identified two distinct IgA1 hinge-region O-glycoform signatures in pediatric IgA vasculitis. The first marked acute disease and was dominated by low sialylated, lower HexNAc-content compositions. The second distinguished IgA vasculitis with renal involvement from IgA vasculitis without renal involvement. The renal signal was independent of total IgA1 abundance and was carried by a small number of glycoforms.

### Total IgA1 abundance

The sequence coverage of IgA1 was important, with seven peptides spread across the protein sequence. Peptide 154-168 (DASGVTFTWTPSSGK) was used to quantify IgA1 and to correct the variation in total IgA1 for the following steps: glycopeptide analysis. Quantity of circulating IgA1 was higher in acute phase of IgAV (A) than patients in remission of IgAV (B) and higher than healthy controls (HCs) (C). Most studies on the pathophysiology of IgAV have reported dysregulation of IgA production with increased serum IgA concentrations [17]. IgA was also quantified routinely and this parameter was present in the patient data. Same tendency was observed with a decrease by 56% between A and B, and 96% between A and C. Total IgA1 levels normalize during remission, consistent with the findings from the O-glycosylation analysis. The total IgA1 levels did not differ according to renal status. Therefore, renal separation reflected glycoform composition rather than IgA1 concentration.

### IgA O-glycosylation

Biochemically, the IgA1 hinge region is uniquely suited to generate disease-relevant heterogeneity because it contains clustered mucin-type O-glycans. High-resolution glycoproteomic work has shown that the IgA1 hinge region includes nine potential Ser/Thr O-glycosylation sites, but typically only three to six O-glycans are occupied, generating a wide distribution of glycoforms. These O-glycans are primarily core 1 structures built stepwise from GalNAc, extended variably by galactose, and capped by sialic acid, creating extensive microheterogeneity in (i) site occupancy, (ii) galactosylation status, and (iii) sialylation patterns [27, 29, 35]. This complexity is central to the concept of Gd-IgA1, where undergalactosylation exposes GalNAc-containing epitopes with potential immunogenicity and altered receptor interactions. However, translating this mechanistic framework into clinically actionable tools has been difficult, in part because hinge-region O-glycosylation is analytically challenging to measure at scale and with site-specific resolution. Mass spectrometry (MS) has become the principal technology for mapping IgA1 hinge glycoforms, including approaches that can assign O-glycosylation sites and quantify isomeric glycoforms. This microheterogeneity creates many isomeric glycopeptides, which remain analytically challenging. The data presented in this study are compositional. H denotes hexose and N denotes HexNAc. These values cannot be directly equated with galactose and GalNAc, and they do not resolve linkage, branching, or site-specific isomers.

Site assignments should remain putative because the fragmentation method has not been optimized for unambiguous localization. Electron-transfer-based fragmentation or orthogonal glycan analysis would strengthen the structural assignments [27, 29].

IgA1 hinge-region O-glycoform abundances between patients in the acute phase of IgA vasculitis (IgAV, A) and patients in remission (B) were compared to identify clinically interpretable glycan biomarkers of disease phase. IgA1 hinge-region O-glycans are predominantly core-1 structures (GalNAc with β1,3-linked galactose) and exhibit variable sialylation. Altered hinge O-glycosylation (including galactose-deficient patterns) has been implicated in the IgA disease spectrum through immune complex formation. Our workflow quantified 69 compositions over a wide dynamic range using only 5 µL of plasma. Duplicate measurements showed a median coefficient of variation of 6%. The analytical depth and sample efficiency are important strengths of this method.

The cohort was dominated by sialylated species and N4–N6 compositions, consistent with the known multiplicity of IgA1 hinge-region O-glycans. The large number of low-abundance species also explains why manual peak review and strict quality criteria were necessary [27, 35, 36].

### A strong acute IgA vasculitis signature

The acute signature was large and internally consistent. Thirty-five glycoforms changed in the same direction in both remission and healthy controls. Several single glycoforms reached AUCs of 0.93–0.95. The biological axis was also clear in this study. Acute IgA vasculitis was enriched in S0–S1 compositions and depleted of highly sialylated HexNAc-rich compositions. The acute-phase group (“A”) exhibited hyposialylation and reduced glycan branching. This profile is characterized by an accumulation of simple agalactosylated and/or asialylated glycans, at the expense of highly sialylated complex glycans, which are conversely more abundant in the control group. Derived traits confirmed this pattern but did not improve the classification. This result favors a parsimonious marker panel and shows that the summary indices capture the same underlying glycosylation axis as the individual markers.

Disease-state analysis identified broad and coordinated remodeling of the IgA1 hinge-region O-glycoform profile during acute IgAV. The inverse relationship between acute-phase enrichment and sialic acid content indicates that this pattern was not driven by a few isolated glycoforms. Instead, it followed a systematic compositional axis dominated by reduced sialylation and, to a lesser extent, by lower HexNAc content. This finding is biologically plausible, given the marked microheterogeneity of the IgA1 hinge region. Up to six clustered O-glycans can be attached to the hinge peptide, with variable site occupancy, galactosylation, and sialylation [27]. Aggregate Gd-IgA1 assays do not resolve this diversity. In contrast, mass spectrometry studies on IgA nephropathy have shown that individual IgA1 hinge glycoforms and derived glycosylation traits contain clinically relevant information. Chen et al. quantified 42 hinge-region O-glycopeptides and reported reduced sialylation in IgA nephropathy, together with an association between sialylation and eGFR [31]. Zhang et al. similarly found lower GalNAc and galactose content in IgA nephropathy than in disease and healthy controls, and showed that composition-derived traits discriminated disease more effectively than global Gd-IgA1 measurements [32].

Our results extend these observations to pediatric IgAV and identify reduced sialylation as a major disease-state-associated dimension of IgA1 remodeling.

### Attenuation of IgA1 hinge-region O-glycoform remodeling in remission

The remission group exhibited a substantially attenuated pattern. Although the direction of the structure– abundance relationship remained aligned with that observed in acute disease, the magnitude of the fold changes was markedly lower and no individual glycoform remained significant after FDR correction. Therefore, the nominal differences observed for several highly sialylated glycoforms should not be interpreted as a validated remission signature. Because the acute and remission groups comprised different children, the data are consistent with a glycoform profile closer to that of healthy controls in remission, but they do not demonstrate within-patient normalization. This distinction is important because circulating Gd-IgA1 has a strong heritable component in pediatric IgA-mediated kidney disease, suggesting that constitutive glycosylation susceptibility may coexist with disease-state-dependent remodeling [15]. Longitudinal mass spectrometry studies in IgA nephropathy have also shown that specific IgA1 glycoforms can change after treatment, supporting the biological plasticity of the glycoform profile; however, comparable longitudinal evidence is not yet available in pediatric IgAV [37].

### Renal-involvement-associated IgA1 O-glycoform signature

Renal comparison is the most clinically relevant contribution. H2N2S1, H3N3S5, H3N4S4, and H4N4S3 distinguished IgA vasculitis with renal involvement from that without renal involvement, with AUCs of 0.75–0.83. The combined score achieved an internally cross-validated AUC of approximately 0.85. This differs from earlier studies, which largely compared IgA vasculitis nephritis with healthy controls, IgA nephropathy, or other end-stage kidney diseases. The direct separation of renal from non-renal pediatric IgAV addresses the actual stratification problem.

Renal analysis was consistent with previous evidence linking abnormal IgA1 O-glycosylation to kidney involvement in IgAV. Allen et al. first reported abnormal lectin binding of circulating IgA1 in patients with clinical nephritis, but not in patients with IgAV without renal involvement [24]. Lau et al. subsequently found elevated serum Gd-IgA1 levels in children with IgAV nephritis compared to healthy children and renal-disease controls [23]. In a prospective pediatric cohort, Pillebout et al. observed higher circulating Gd-IgA1 in IgAV with nephritis than in IgAV without nephritis and reported an AUC of 0.73 for the renal-status comparison [26]. However, the results have not been fully consistent across cohorts. In a larger Chinese pediatric study, Gd-IgA1 was clearly elevated in IgAV nephritis relative to healthy controls, whereas its difference from IgAV without nephritis did not reach statistical significance [25]. Mechanistic studies have supported the renal relevance of this pathway. Suzuki et al. found increased production of both Gd-IgA1 and Gd-IgA1-specific IgG autoantibodies in IgAV nephritis, together with altered expression of enzymes involved in IgA1 O-glycan biosynthesis [38]. In adults with IgAV nephritis, higher plasma Gd-IgA1 levels were independently associated with subsequent loss of kidney function [39]. Collectively, these studies support a relationship between aberrant IgA1 glycosylation and renal disease, but they also show that a single aggregate Gd-IgA1 measurement provides only moderate and variable discrimination between IgAV with and without nephritis.

Our composition-resolved analysis refines the findings of previous studies. A broad inverse relationship between glycoform abundance and sialic acid count was observed in both IgAV groups relative to healthy controls. Therefore, reduced sialylation should be interpreted as a general IgAV-associated axis rather than a renal-specific abnormality. Renal involvement was associated with a smaller set of individual compositions, particularly H2N2S1, H3N3S5, H3N4S4, and H4N4S3. These glycoforms differed between IgAV with and without nephritis but showed little or no separation between healthy controls and non-nephritic IgAV patients. This pattern supports a renal status association that is distinct from broader disease-associated remodeling. Nakazawa et al. previously used MALDI-TOF mass spectrometry in seven adult transplant recipients with end-stage IgAV nephritis and found lower GalNAc content and reduced galactose attachment, with profiles resembling those of IgA nephropathy [30]. This study was limited to advanced adult kidney disease and used a workflow that did not retain the sialylation dimension. The present study extends this work by profiling intact sialylated compositions in children and directly comparing IgAV with and without renal involvement. The combined renal score achieved an AUC of 0.86 (IC 95 % 0.7–0.94) in the present cohort. This value is promising but should not be interpreted as superior to the AUC of 0.73 reported for Gd-IgA1 by Pillebout et al., because the cohorts, assays, endpoints, marker-selection procedures, and validation strategies differed.

A potential concern was that the pooled comparison according to renal status included both acute-phase and remission samples and could therefore have been influenced by disease activity. However, acute and remission samples were similarly distributed between the renal-status groups. Moreover, all four glycoforms were less abundant with renal involvement in both phases, with no statistical evidence of phase-by-renal-status interaction. These observations support an association with concurrent renal status that is not solely explained by disease activity. Because the subgroup sizes were small and the markers were selected in the same cohort, the absence of a detected interaction should not be interpreted as evidence of equivalent effects across phases. Independent longitudinal validation remains required.

### Strengths and limitations

Key strengths include the exclusively pediatric cohort, the low plasma input volume (5 µL), preservation of intact sialylated glycoforms, quantification of 69 IgA1 hinge-region O-glycoform compositions, good analytical repeatability in duplicate measurements (median CV, 6%), correction for multiple testing, and direct comparison of children with IgA vasculitis with versus without concurrent renal involvement. Limitations include the modest sample size including possible clinical and batch confounding, and limited structural resolution of glycan isomers and sites.

## CONCLUSIONS

Pediatric IgA vasculitis is associated with a strong IgA1 hinge-region O-glycoform signature characterized by a reduction in sialylation, which is a broad IgAV-associated feature, whereas renal involvement is associated with a smaller and composition-specific IgA1 O-glycoform panel. These findings extend previous lectin-based and desialylated MALDI-TOF studies by preserving sialylation and directly addressing renal stratification in children. Importantly, these renal-associated changes were observed both during the acute phase and in remission, indicating that they reflect renal involvement rather than disease activity. However, independent and longitudinal validation is required before clinical use.

### Perspectives

Prospective sampling from diagnosis through follow-up should be performed to test whether the renal-associated panel exists before clinical nephritis. The primary endpoint should be incident renal involvement, defined by standardized urine, blood pressure, kidney function, and biopsy criteria.

A validated assay could eventually complement the routine monitoring. The present data do not justify the altered biopsy thresholds or presymptomatic corticosteroid treatment.

## Supporting information

Supplementary Data 1. Internal validation of the four-glycan classifier

Supplementary Table 1. Identified IgA1 hinge-region O-glycopeptides

Supplementary Table 2. Technical repeatability of IgA1 O-glycopeptide quantification

Supplementary Table 3. Raw Skyline peak areas

Supplementary Table 4. Normalized IgA1 O-glycopeptide peak areas

Supplementary Table 5. Differential analyses of IgA1 O-glycopeptides

Supplementary Table 6. Study population

## Data Availability

All data produced in the present study are available upon reasonable request to the authors

## List of abbreviations

AUC: area under the receiver operating characteristic curve
FDR: false discovery rate
Gd-IgA1: galactose-deficient immunoglobulin A1
IgAN: immunoglobulin A nephropathy
IgAV: immunoglobulin A vasculitis
IgAVN: immunoglobulin A vasculitis with nephritis
LC–MS/MS: liquid chromatography–tandem mass spectrometry

## Declarations

### Ethics approval and consent to participate

The FOX-TREG study and the subsequent use of archived biological samples were approved by the CPP Sud Méditerranée III ethics committee (approval no. 2013.10.05) and by the relevant institutional review board under approval nos. 19.01.06 and 20.0061. The associated biological collection was declared under reference DC-2019-3463. Written informed consent was obtained from participants’ parents or legal guardians, and age-appropriate information was provided to the children; assent was obtained whenever applicable.

### Consent for publication

Not applicable.

### Availability of data and materials

The mass spectrometry data and Skyline documents generated in this study have been deposited in Panorama Public through the ProteomeXchange Consortium under dataset identifier PXD082508 and Panorama Public DOI 10.6069/XXXX-XXXX. The associated quantitative data are provided in the Supplementary Information.

### Authors’ contributions

JV analysed the results and drafted the manuscript. CH, JV, PC, TTA and AF conceived the study. CH and JV supervised the LC–MS analyses, analysed the results, and contributed to drafting the manuscript. AF participated to the first draft of the article. RC performed the pre-analytic experiments. JK performed the experiments and analysed the data. TC, CR, and MP performed the statistical analyses. AF, TTA, MF, and AS were clinical investigators. All authors read and approved the final manuscript and agreed to be accountable for all aspects of the work.

## Acknowledgements

We thank the patients for their participation. We thank the Centre de ressources biologiques (BB-0033-00032), CHU de Nîmes, groupe hospitalo-universitaire de Carémeau, 30029 Nîmes cedex 09, France. Mass spectrometry experiments were carried out using the facilities of the Montpellier Proteomics Platform (PPM, BioCampus Montpellier), a member of the national Proteomics French Infrastructure (ProFI UAR 2048) supported by the French National Research Agency (ANR-24-INBS-0015, Investments for the future F2030).

---

Supplementary Table 1. Identified IgA1 hinge-region O-glycopeptides and associated mass-spectrometry identification and glycosite-localization metrics.

Supplementary Table 2. Technical repeatability of IgA1 hinge-region O-glycopeptide quantification, summarized by the median coefficient of variation for each glycoform.

Supplementary Table 3. Raw Skyline peak areas of IgA1 hinge-region O-glycopeptides for individual LC–MS runs.

Supplementary Table 4. Median normalized peak areas of IgA1 hinge-region O-glycopeptides for each study participant.

Supplementary Table 5. Univariate and multivariate analyses of IgA1 hinge-region O-glycopeptide profiles according to clinical condition and renal involvement.

Supplementary Table 6. List of patients and healthy controls included in the study population.

**Supplementary Figure 1.** IgA1 hinge O-glycosylation - composition & site-localization. A: Rank-abundance of the O-glycan profile. Median % of total HNS for each of the 69 glycans, ranked from most to least abundant (log-scaled y-axis). The accompanying single-column list gives the rank order of every glycan. B: HexNAc-Hex imbalance (GDI). Median % of total HNS grouped by the galactosylation-deficient index GDI = N - H (excess of HexNAc over hexose). C: Sialylation degree. Median % of total HNS carried by glycans grouped by their number of sialic-acid residues (S = 0-5). D: ‘Occupancy/complexity’ proxy by HexNAc count. Median % of total HNS grouped by the number of HexNAc residues (N = 1-6). E: Weighted localization probability across the peptide sequence. Best-position localization were extracted from peptide-spectrum matches corresponding to the peptide HYTNPSQDVTVPCPVPSTPPTPSPSTPPTPSPSCCHPR (positions noted on the x-axis, 1-indexed). The heatmap color intensity represents this normalized weighted probability at each residue; amino-acid letters and residue indices are overlaid for sequence context.

**Supplementary Figure 2.** Renal-associated changes in the four IgA1 O-glycoforms are independent of disease activity. For each marker, mean log2 abundance (+/- 95% CI, individual samples shown as faded points) is plotted against renal status, separately for patients in acute (A, purple) and in remission (B, green). The two lines decline in parallel, indicating that nephritis lowers each glycoform by a similar amount irrespective of disease activity; the activity x renal interaction was not significant for any marker (FDR 0.36-0.76, two-way model on log2 abundance).

