## Supplementary Data 1. Internal validation of the four-glycan classifier for "IgA1 hinge-region O-glycoform signatures associated with disease phase and kidney involvement in pediatric IgA vasculitis: a cross-sectional mass-spectrometry study"

The ability of the four-glycoform panel to separate IgAV patients with and without renal involvement was assessed by repeated stratified five-fold cross-validation (200 repeats of the partition). Two quantities were estimated. First, to evaluate the complete discovery procedure without information leaking from the held-out patients, glycan ranking (two-sided Mann-Whitney), selection of the four most discriminant glycans, z-score standardisation and logistic regression were all performed within each training fold, and the held-out patients were scored. Second, for reference, the pre-specified panel was cross-validated with the selection step left unchanged, an estimate that remains optimistically biased because the panel was defined on the whole cohort. Predicted probabilities were averaged over repeats, and discrimination was summarised by the AUC with a 95% confidence interval from 3,000 patient bootstrap resamples; the dispersion of the estimate across random partitions is reported separately and is not a confidence interval. Calibration was assessed by unpenalised logistic recalibration of the out-of-sample probabilities (slope and intercept, with bootstrap confidence interval) and by the Brier score. The stability of the selection step was quantified as the percentage of training folds in which each glycan entered the top four.

### Results

When the entire discovery procedure - ranking, selection of four glycans, and model fitting - was repeated inside each training fold, the resulting classifier separated nephritis from no-nephritis with an out-of-sample AUC of 0.86 (95% CI 0.75-0.94; Figure 1A). The apparent AUC obtained by fitting and evaluating the pre-specified panel on all patients was 0.91, so the total optimism of the naïve estimate was 0.05 (Figure 1B). The selection step was highly reproducible: over the 1,000 training folds, H2N2S1 was reselected in 100% of folds, H3N3S5 and H3N4S4 in 94%, and H4N4S3 in 66%, the only other frequently chosen glycan being H3N5S4 (32%; Figure 1D), indicating that the panel reported here is not an unstable artefact of a single partition of the data. Out-of-sample probabilities showed no evidence of systematic miscalibration (slope 0.99, intercept -0.01, Brier 0.16), although the slope was estimated imprecisely in this cohort size (95% CI 0.64-1.94; Figure 1C).

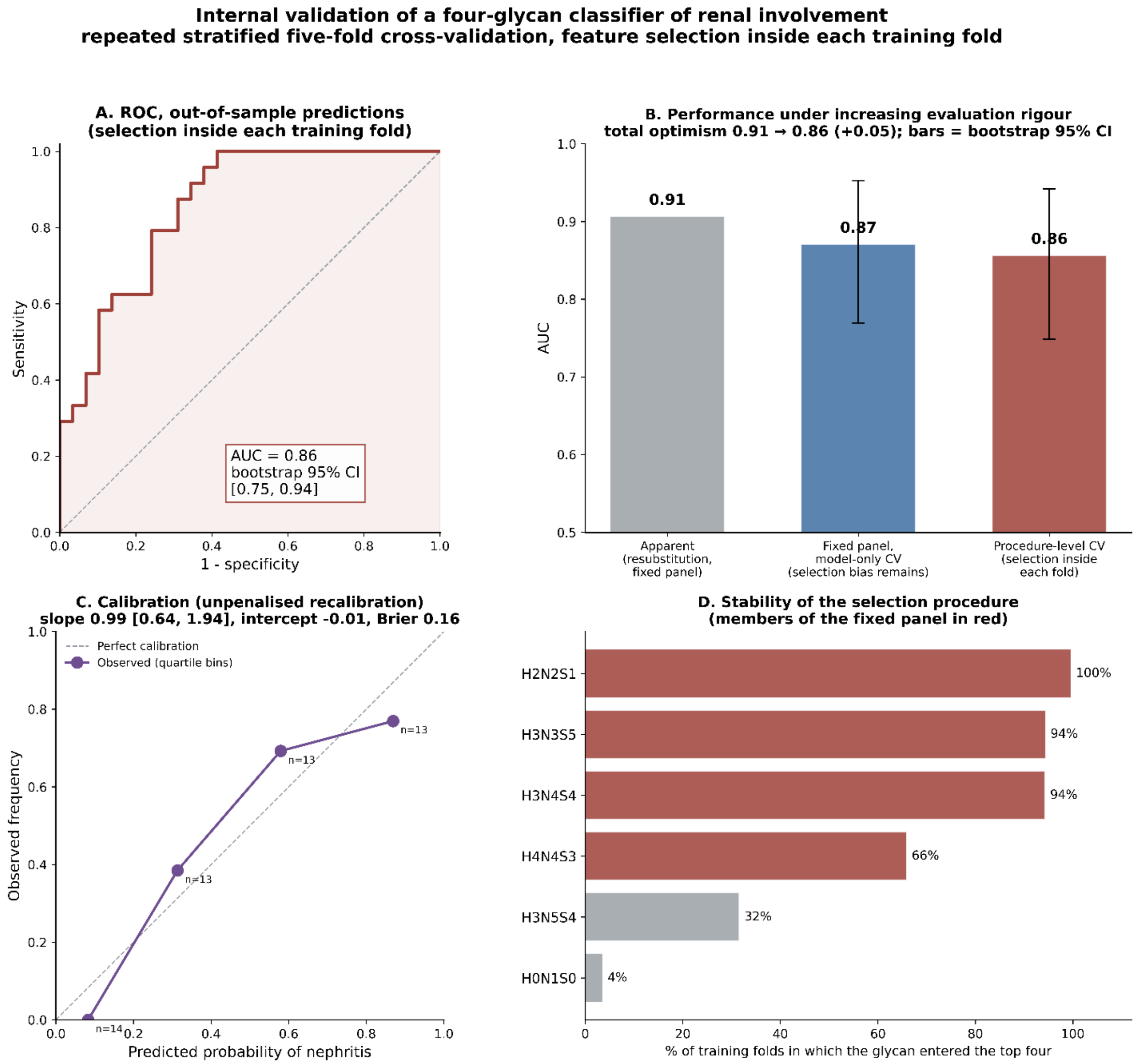


Figure 1. Internal validation of a four-glycan classifier of renal involvement by repeated stratified five-fold cross-validation with feature selection performed within each training fold. (A) ROC built from out-of-sample predicted probabilities averaged over 200 repeats of the partition; the AUC is given with its patient-bootstrap 95% confidence interval. (B) AUC under three levels of evaluation rigour: apparent (the pre-specified panel fitted and evaluated on all patients), cross-validation of the model with the panel held fixed (the selection step still used every patient, so this remains optimistically biased), and

cross-validation of the complete procedure with selection repeated inside each training fold; error bars are patient-bootstrap 95% confidence intervals. (C) Calibration of the out-of-sample probabilities in quartile bins, with the slope and intercept from an unpenalised logistic recalibration and the Brier score. (D) Stability of the selection step: percentage of the 1,000 training folds in which each glycan entered the top four; members of the pre-specified panel are shown in red. Note that (A), (C) and (D) characterise the discovery procedure and not the fixed panel, whose unbiased evaluation requires an independent cohort.
