## Supplementary figures and images for "IgA1 hinge-region O-glycoform signatures associated with disease phase and kidney involvement in pediatric IgA vasculitis: a cross-sectional mass-spectrometry study"

### Supplementary Figures

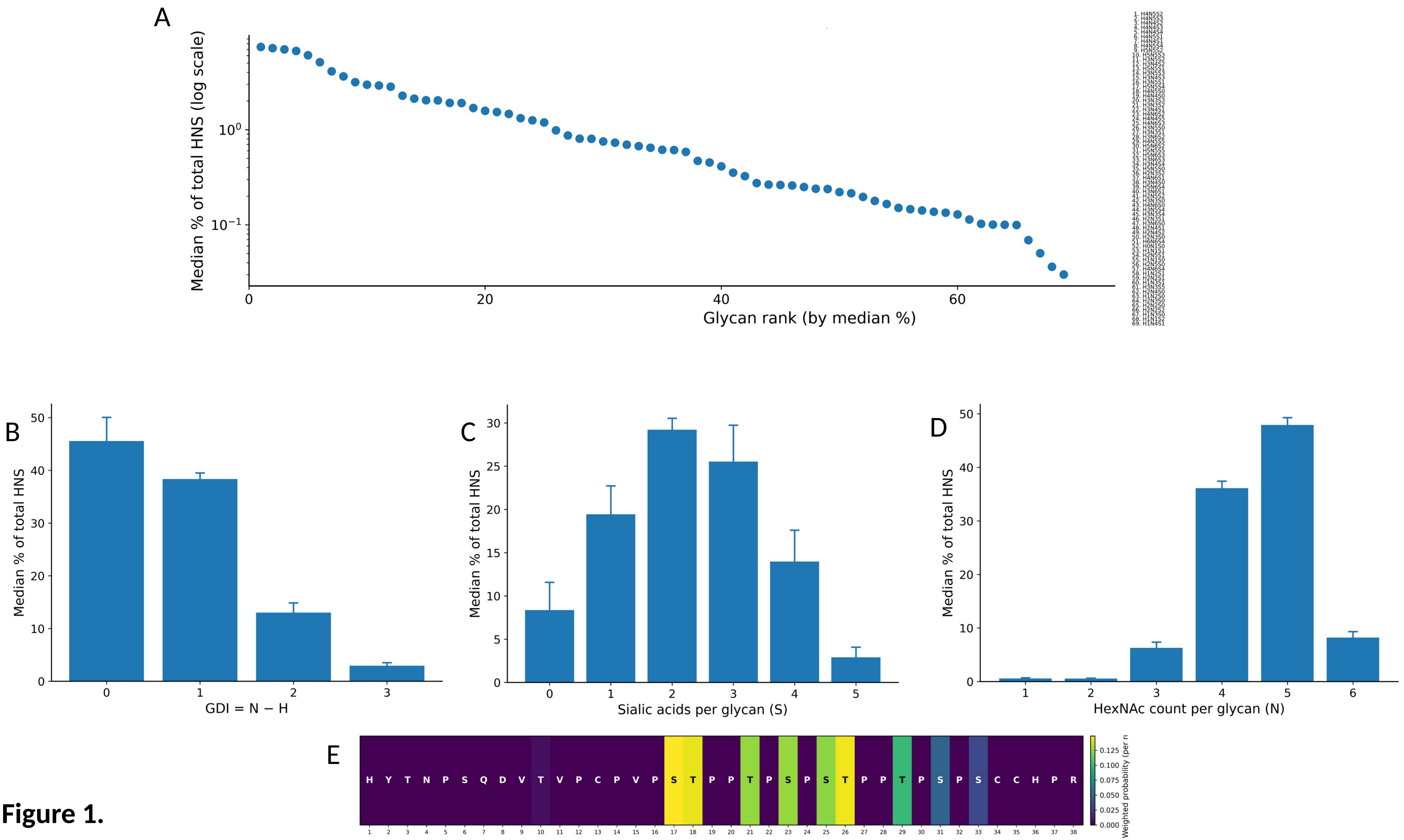

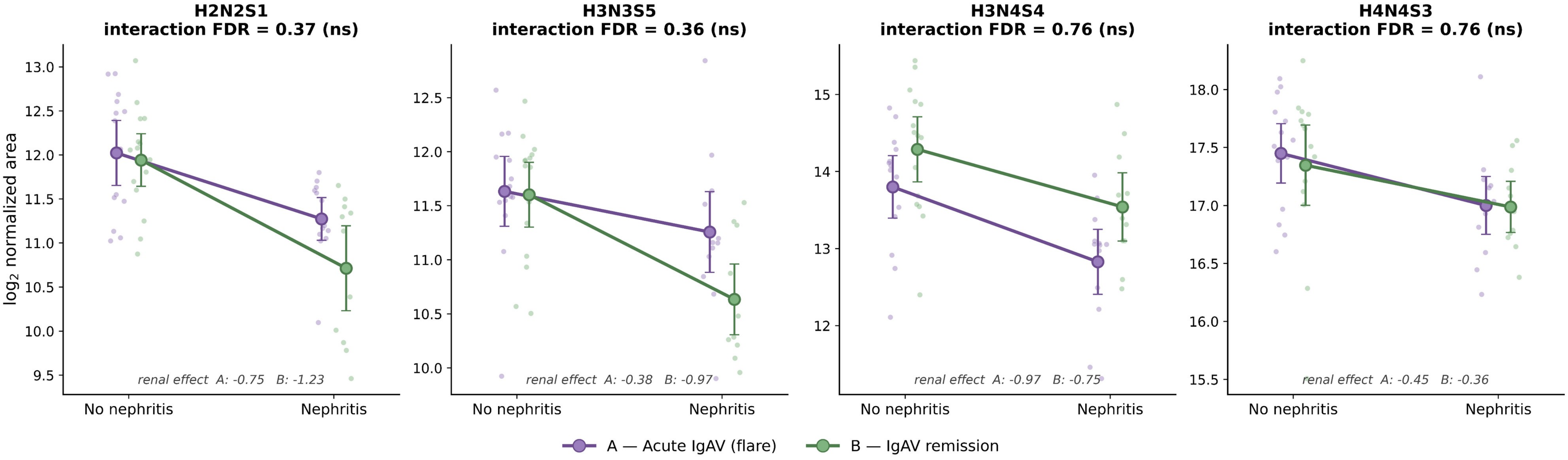
